# British Oncology Pharmacy Association Delphi Consensus Guidelines: co-infusion of trometamol-containing calcium folinate (Leucovorin) with Systemic Anti-Cancer Treatments

**DOI:** 10.1101/2024.02.10.24302100

**Authors:** Calum Polwart, Tim Root, Songül Tezcan, Sharon Meehan, Bill Wetherill, Chloë Waterson, Bruce Burnett, Rena Chauhan, Ibrahim Al-Modaris, Rhiannon Walters-Davies

## Abstract

Drug stability and compatibility are critical factors influencing cost and logistics of treatment delivery, therapeutic effectiveness, and patient safety. This is particularly significant in the realm of cancer chemotherapeutics, where stability and compatibility studies play a vital role in ensuring rational and safe medicine administration. Oxaliplatin, fluorouracil, and irinotecan, commonly used in various combination for gastrointestinal cancers, are complemented by co-administration of folinic acid in certain protocols. Notably, some folinic acid preparations include trometamol as an excipient, potentially impacting the stability of the chemotherapeutic agentsif infused concomitantly. This study seeks to establish guidelines for oncology multidisciplinary teams, addressing potential risks associated with the combination of trometamol-containing folinic acid and chemotherapeutics. To achieve this, a quantitative questionnaire was distributed to members of the British Oncology Pharmacy Association (BOPA) and non-BOPA members through an online survey. Nineteen healthcare professionals with oncology experience, comprising 18 pharmacists and 1 nurse, completed the questionnaires. Each participant rated the validity and clarity of statements on a 5-point scale. The Delphi process concluded after the fourth round, consolidating the findings and recommendations from the multidisciplinary team. Twelve recommendations for safe practice have been made.

## Introduction

Folinic acid (Leucovorin) in combination with fluorouracil has been the backbone of cytotoxic chemotherapy for colorectal and many other gastrointestinal cancers for almost four decades^1^. In the United Kingdom (UK), the most used regimens are an adaptation of the de Gramont regimens^2^, using co-administration of oxaliplatin and/or irinotecan sometimes in combination with a targeted antibody^3^. These regimens conventionally give doses of at least 200 mg/m^2^ folinic acid as a two-hour infusion immediately prior to commencing the 46-hour fluorouracil infusion. Dosing of folinic acid varies considerably between protocols and institutions^1^. When administered alongside other cytotoxic chemotherapy, folinic acid is usually administered through the same single lumen intravenous line using a Y-line administration set. Folinic acid is usually administered as the calcium salt (calcium folinate), although a sodium folinate preparation is also available. In North America, folinic acid is referred to by its original brand name Leucovorin^4^. Only the *levo-*isomer is considered clinically active, and products containing either *levo-*folinate and a racemic mixture are available^4^.

The Manufacturing Authorisation Holder’s (MAH) Statement of Product Characteristics (SmPC) for oxaliplatin states: “*Oxaliplatin 85 mg/m^2^ intravenous infusion in 250 to 500 ml of glucose 5 % (50 mg/ml) solution is given at the same time as folinic acid (FA) intravenous infusion in glucose 5 % solution, over 2 to 6 hours, using a Y-line placed immediately before the site of infusion.”*^5^ However, it then states: “*These two medicinal products should not be combined in the same infusion bag. Folinic acid (FA) must not contain trometamol as an excipient and must only be diluted using isotonic glucose 5 % solution, never in alkaline solutions or sodium chloride or chloride containing solutions.*”^5^. The basis for this statement and the clinical significance of the implied chemical incompatibility is unclear, even after correspondence with the marketing authorisation holder. Until recently, this has not posed a problem in the UK, as no folinic acid preparation contained trometamol as an excipient. However, a product is now available in the UK which contains 10mg/mL of trometamol^3^.

Trometamol-free folinic acid has been subject to UK and international supply shortages on several occasions^7^, and there has been insufficient supply of alternative trometamol-free calcium folinate products. This has meant that some UK hospitals have had no option but to use a trometamol-containing product with a lack of consensus and variety of approaches to mitigation of the risk of incompatibility and as such guidance surrounding its safe implementation may be beneficial.

## Objectives

We sought to create clinical practice guidelines for British Oncology Pharmacy practitioners and other members of the UK oncology multidisciplinary team to address the potential chemical and clinical significance of the risks associated with combination of a trometamol-containing folinic acid with cytotoxic chemotherapy.

## Methodology

The British Oncology Pharmacy Association’s (BOPA) on-line discussion forum was used as a platform to gauge interest, and selected parties were subsequently assembled. A comprehensive literature search had been completed to establish the evidence for, and severity of the potential interaction with trometamol. Due to the lack of published literature that confirmed the basis for, or significance of a potential interaction with trometamol, we started to develop a modified Delphi-method^8^ in order to reach consensus on recommendations for safe practice. The steps involved in this process are described in Figure 1. Recruitment for a second and third round of Delphi voting was widened to the entire BOPA membership (via email), and non-BOPA members through social media and email, and through the NHS Pharmaceutical Quality Assurance network. Following the first round of discussion, new evidence from a Canadian study^9^ was published which suggested the combination was unlikely to have any adverse effect on efficacy or toxicity.

**Figure 1:**
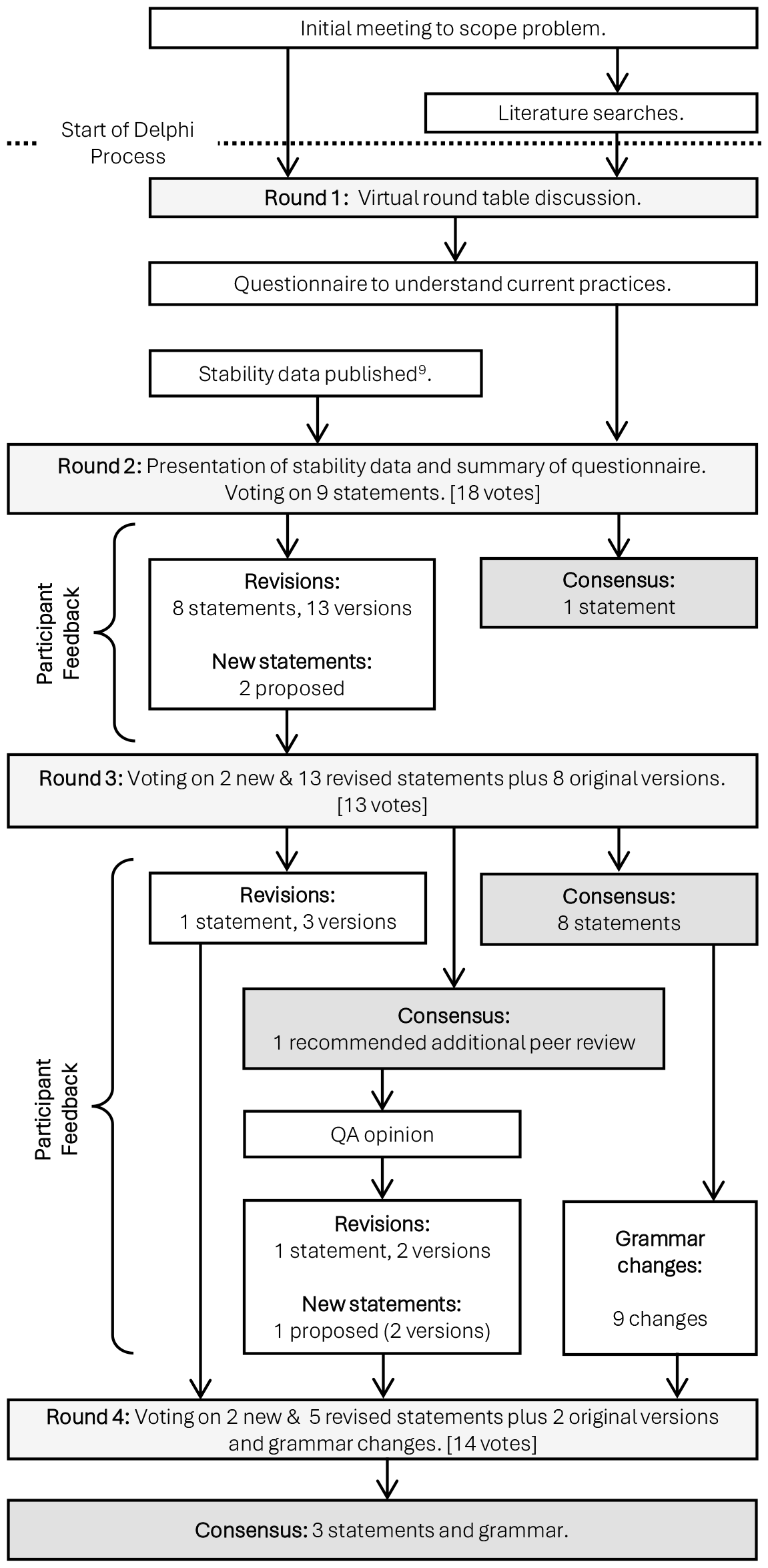
Stages of modified Delphi process. Details of statements are in the supplementary materials. A total of 12 statements reached consensus. No statements without an alternative failed to reach consensus.

Membership of the first, and second to fourth round panels is described in the supplementary material. Representation at the voting rounds included: 18 pharmacists and 1 chemotherapy nurse. 11 of the pharmacists stated they had expertise in Upper GI cancers, 12 in Lower GI cancers, three with wider clinical oncology experience, and seven with aseptic expertise; in addition to one each with, medicines procurement, pharmaceutical quality assurance, academic and specialist pharmacy support services. A questionnaire was not used in the first round. This was conducted through an online open table discussion which was helpful to set the premise for further rounds and was followed by a quantitative questionnaire distributed to BOPA members by email and social media. The survey aimed to understand variations in established practice. The results were summarised in the research protocol which was shared with all panellists prior to a second online meeting^10^.

Immediately prior to round two, the panellists participated in an online oral presentation, summarising the evidence to date including a presentation from the author of the Canadian Study^9^. From this, there posed an opportunity to ask any questions. All voting and suggestions for statement revision were then collated and fully anonymised using an on-line form. Voting used a Likert scale; (1 = Strongly Agree, 2 = Agree, 3 = Neither Agree nor Disagree, 4 = Disagree, 5 = Strongly Disagree). Consensus agreements was defined as when the median score was identified as 2 or less (i.e. at least half the participants agree or strongly agree with a statement) and the inter-quartile range (IQR) of responses was identified as 1.5 or less. (i.e. at least half the participants score within 1 point of each other). Consensus disagreement would be reached for a median score of 4 or more, with the same IQR. If neither was reached, then no formal consensus had been reached.

When multiple iterations of the same statement were considered, the score with the lowest median score and an IQR ≤1.5 was deemed to be the preferred option. A maximum of three rounds of voting were permitted. The original intention was to undertake two or three rounds of voting in the same evening, which would take place directly following the meeting. No threshold for voting was defined in advance as it was not expected there would be a loss of panel members to follow-up but it soon became clear that further rounds on the same evening was not practicable. We therefore adopted the conventional threshold of 70% of original voting panellists needing to respond to subsequent voting rounds or further votes would not be held. Reminder emails were sent to all participants to further prompt voting.

## Funding

This project has not been subject to external funding. No conflicts of interest were declared by any of the panel members or authors.

### Target users

Our consensus is intended to be useful for pharmacist and non-pharmacy clinicians, pharmacy technicians and other healthcare professionals practicing oncology pharmacy in the UK, but is likely to be useful to others internationally where calcium folinate products containing trometamol are available.

### Search methods

We searched for compatibility data on Medline (via PubMed), EmBase, CINAHL and International Pharmaceutical Abstracts using the search terms:

- “oxaliplatin” OR “trometamol” AND
- “calcium folinate” OR “calcium levo-folinate” OR “folinic acid” OR “levo-folinic acid” OR “sodium folinate” OR “sodium levo-folinate” OR “leucovorin” OR “levo-leukovorin” AND
- “compatibility” OR “incompatibility” OR “stability” OR “instability” OR “precipitation”.

## Formulation of recommendations

Following the presentation of evidence, the panel initially voted on nine statements (listed in the Supplementary Material). All statements were considered acceptable, however, panellists made recommendations to amend eight of the nine statements. Two additional statements were also proposed. Panellists were then asked to vote on the original eight statements, two new statements and modified versions of the original eight statements.

Following the second cycle of voting, consensus was reached on ten of the eleven statements. However, one statement about which there was greatest consensus specified that further peer review of the Canadian study^9^ should take place. We therefore sought peer review from colleagues in the NHS Specialist Pharmacy Service Quality Assurance team.

They kindly provided an opinion about the poster and an overview of the facts as they were known surrounding trometamol compatibility with oxaliplatin and the oxaliplatin degradation pathway. The opinion was shared with Delphi panellists by email, with a summary of the second round of voting. Participants were asked to suggest revised wording (by email response to the lead author) for the disputed statement.

Some participants had also highlighted grammatical errors in statements and general improvements to enhance inclusivity and robustness of resulting recommendations. Participants were then asked to vote for a final round on the revised statements and to accept the grammatical modifications.

The recommendations are summarised in

Table 1, and further details of the decision-making process are provided in the following text.

**Table 1.**
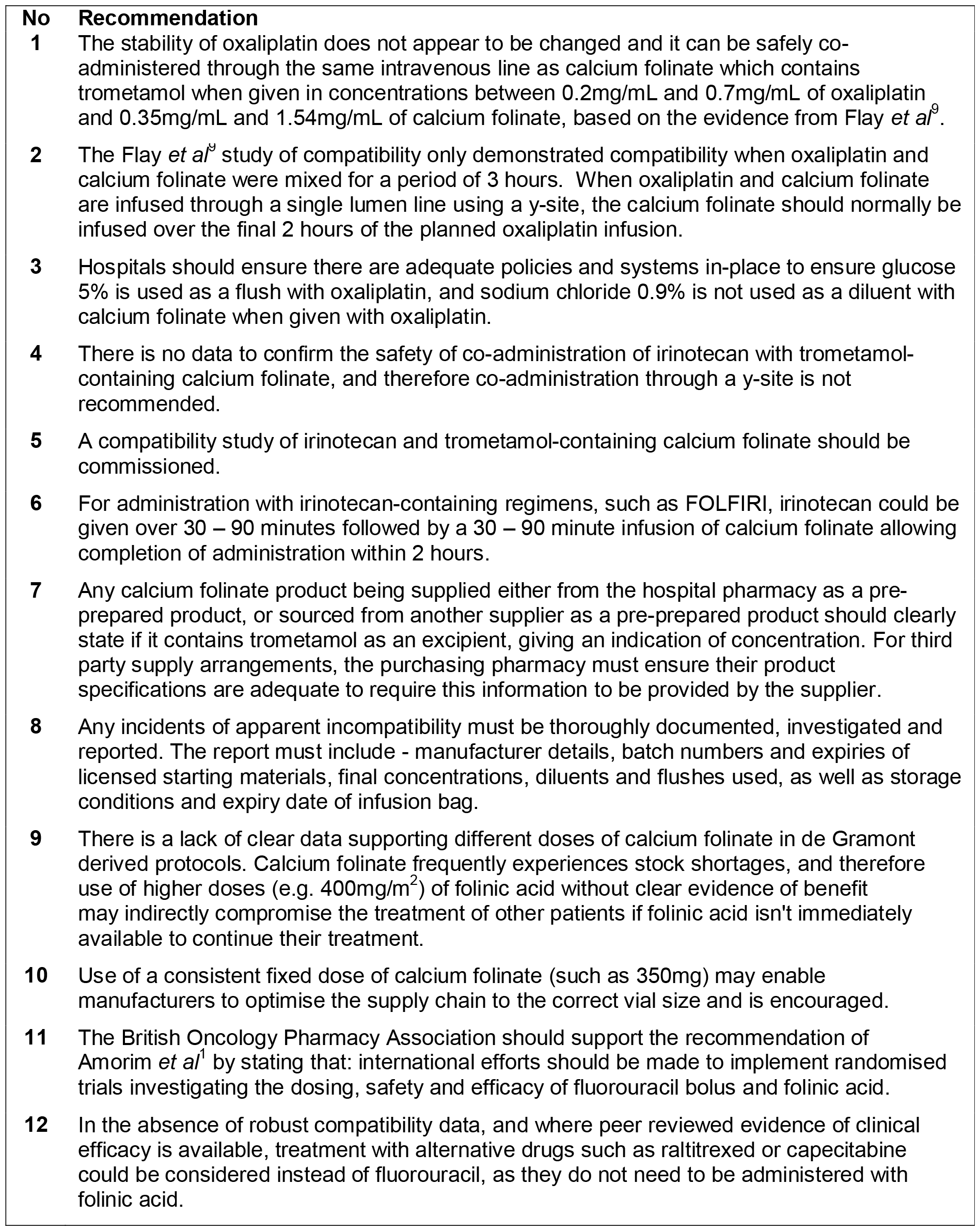
Summary of recommendations from the Delphi Panelists.

### Compatibility with oxaliplatin

Oxaliplatin is given by intravenous (IV) infusion over two to six hours, and folinic acid has usually been administered at the same time, through the same IV access via a Y-type giving set, at the same rate. This is more efficient for the service, on daycare ambulatory chemotherapy units which is a recognised challenge^11^, and a better experience for the patient as less time is required for treatment. Any change in practice which increases the time a patient is on the unit decreases overall capacity. A potentially substantial inconvenience to patients is likely; some will be travelling some distance for treatment on a two-weekly schedule for sometimes more than a year. However, an incompatibility of trometamol with oxaliplatin might also create clinical risks for patients – either from loss of oxaliplatin and/or folinic acid potency which could affect efficacy and outcomes, or from harm caused by precipitation. Calcium folinate potentiates the effectiveness of fluorouracil hence non-availability of trometamol-free calcium folinate might also pose risks of harm for patients.

Prior to publication of the Canadian study^9^, much of the discussion at the round table meeting had been focused on finding alternative mitigations to co-administration via a Y-site such as use of double lumen lines or placement of a second peripheral line. However, the new data^9^ provided no evidence that the presence of trometamol was problematic when using Y-site administration. The panel therefore reviewed several statements, and having sought additional peer review of the study, concluded unanimously agreed (64% strongly agree, IQR: 1):

> ***Recommendation 1:***
>
> *The stability of oxaliplatin does not appear to be changed and it can be safely co-administered through the same intravenous line as calcium folinate which contains trometamol when given in concentrations between 0.2mg/mL and 0.7mg/mL of oxaliplatin and 0.35mg/mL and 1.54mg/mL of calcium folinate, based on the evidence from Flay et al^9^.*

The concentration ranges included in this recommendation differ slightly from established UK practice but are based on the parameters of the stability study; it is expected that some UK centres would need to update their standard concentration limits to match these parameters. The compatibility study was based on in vitro mixing of oxaliplatin and folinic acid with trometamol for a period of up to three hours. The peer review of the study noted that in clinical practice oxaliplatin could be administered over periods of up to six hours.

Panellists were therefore asked to consider two versions of a further statement, one recommending “calcium folinate should normally be infused over the final two hours of the planned oxaliplatin infusion” and one which considered that because the contact time in the Y-site was short there was no concern by the three hour limitation of the study. The first of these alternative statements was preferred and reached consensus (77 % agree/strongly agree, median score: 2, IQR: 0.75).

> ***Recommendation 2:***
>
> *The Flay et al^9^ study of compatibility only demonstrated compatibility when oxaliplatin and calcium folinate were mixed for a period of 3 hours. When oxaliplatin and calcium folinate are infused through a single lumen line using a y-site, the calcium folinate should normally be infused over the final 2 hours of the planned oxaliplatin infusion.*

The survey of UK practice identified that 19% of respondents did not specify that glucose 5% injection must be used to flush the line before and after oxaliplatin infusion. Routine practice for flushing IV lines would be to use sodium chloride 0.9%, which is known to be incompatible with oxaliplatin^5^. During the first round of voting panellists strongly agreed (Median score: 1, IQR: 0) a statement about the need to use glucose as a flush and diluent. After minor grammatical revisions this was modified to:

> ***Recommendation 3:***
>
> *Hospitals should ensure there are adequate policies and systems in-place to ensure glucose 5% is used as a flush with oxaliplatin, and sodium chloride 0.9% is not used as a diluent with calcium folinate when given with oxaliplatin.*

While our recommendation states this applies to hospitals, the advice would equally apply to any healthcare setting where oxaliplatin is being co-administered with calcium folinate.

### Compatibility with irinotecan

When given as part of the FOLFIRI or ‘Irinotecan Modified de Gramont’ regimens, practice for co-administration of calcium folinate is more varied than with co-administration of oxaliplatin. 53% of respondents to our survey indicated they administered treatment concurrently via the same line. Irinotecan can however be given more quickly than oxaliplatin, so sequential treatment poses less of a challenge to service capacity and patient experience.

We are aware of a single study of compatibility of irinotecan with leucovorin^12^, and further details of the excipients are not available. The panellists considered this evidence, and the lack of clear evidence that any incompatibility events had occurred since trometamol-containing calcium folinate has been in use. They considered statements both in favour of and against Y-Site administration in this circumstance and the strongest consensus (Median score 2, IQR 1.5) was:

> ***Recommendation 4:***
>
> *There is no data to confirm the safety of co-administration of irinotecan with trometamol-containing calcium folinate, and therefore coadministration through a y-site is not recommended.*
>
> Furthermore, panellists recommended (median score: 1, IQR: 1) that:

> ***Recommendation 5:***
>
> *A compatibility study of irinotecan and trometamol-containing calcium folinate should be commissioned.*

In an effort to mitigate the adverse impact on service capacity of potential sequential administration of chemotherapy followed by calcium folinate, we sought to confirm if there was an evidence base for administration calcium folinate slowly as in established normal UK practice. We found no clear evidence. The UK Summary of Product Characteristics (SmPC)^6^ states: “no more than 160mg of calcium folinate should be injected per minute due to the calcium content of the solution”. Even for the highest doses of calcium folinate (400mg/m^2^) this would allow administration over as little as six minutes even in larger weight extremes seen in clinically obese patients.

Irinotecan can be given by intravenous infusion over 30 to 90 minutes. The panellists therefore considered several versions of wording which would allow administration of irinotecan followed by calcium folinate in a total time of two hours (the same as when oxaliplatin is administered). They reached the strongest consensus (Median score 2, IQR 0.5) for this statement:

> ***Recommendation 6:***
>
> *For administration with irinotecan-containing regimens, such as FOLFIRI, irinotecan could be given over 30 – 90 minutes followed by a 30 – 90 minute infusion of calcium folinate allowing completion of administration within 2 hours.*

## Product labelling

Our questionnaire revealed that a small number of centres used a third-party pharmacy to prepare their calcium folinate infusions. They indicated that they did not know which brand of calcium folinate they were receiving, and therefore had no knowledge if this contained trometamol as an excipient. Panellists felt that if trometamol was included as an excipient, this should always appear on the label given the explicit contraindication in the Summary of Product Characteristics (SmPC) for oxaliplatin. Therefore, the panellists adopted the following statement (Median score 1, IQR 0):

> ***Recommendation 7:***
>
> *Any calcium folinate product being supplied either from the hospital pharmacy as a pre-prepared product, or sourced from another supplier as a pre-prepared product should clearly state if it contains trometamol as an excipient, giving an indication of concentration. For third party supply arrangements, the purchasing pharmacy must ensure their product specifications are adequate to require this information to be provided by the supplier.*

### Investigation of incompatibility

The panellists felt there should be a systematic approach to investigation of reported incompatibility. During this work, a small number of anecdotal incidents were reported, however, such historical events (some of which occurred prior to availabilty of a trometamol-containing calcium folinate in the UK) lacked sufficient detail to allow assessment of their significance. In some cases incompatibility appeared to be unconnected to the calcium folinate preparation used.

The panellists therefore adopted a recommendation (Median score: 1, IQR: 0) relating to incident investigation, and after grammatical adjustments this reads:

> ***Recommendation 8:***
>
> *Any incidents of apparent incompatibility must be thoroughly documented, investigated and reported. The report must include - manufacturer details,*

*batch numbers and expiries of licensed starting materials, final concentrations, diluents and flushes used, as well as storage conditions and expiry date of infusion bag.*

### Maintaining a resilient supply chain of calcium folinate

There have been numerous occasions where there has been a complete lack of, or very limited supply of calcium folinate available in the UK and elsewhere in the world^7^. Panellists therefore considered if there was anything that could be done to mitigate potential shortages and maintain a resilient supply chain. They questioned the evidence for the vastly varying doses of folinic acid and noted the comments of Amorim and Peixeto^1^ who concluded that fluorouracil bolus and calcium folinate “do not seem to add activity, while it increases toxicity” and recommended “international efforts should be made to implement randomized trials investigating this”.

While there is often a reluctance to change long established practice in a particular setting, the panellists were also concerned that un-evidenced use of high doses of calcium folinate could deplete stocks and deprive some patients of treatment. They therefore adopted two statements (Median Score 1, IQR: 0, and Median Score 1, IQR: 0) regarding preservation and optimisation of supply and dosing:

> ***Recommendation 9:***
>
> *There is a lack of clear data supporting different doses of calcium folinate in de Gramont derived protocols. Calcium folinate frequently experiences stock shortages, and therefore use of higher doses (e.g. 400mg/m^2^) of folinic acid without clear evidence of benefit may indirectly compromise the treatment of other patients if folinic acid isn’t immediately available to continue their treatment.*

> ***Recommendation 10:***
>
> *Use of a consistent fixed dose of calcium folinate (such as 350mg) may enable manufacturers to optimise the supply chain to the correct vial size and is encouraged.*

The panellists also considered a version of recommendation 10 using 300mg of calcium folinate as an example dose, however this was less favoured (Median Score 2, IQR: 1.75). It should also be noted that there is a suggestion that higher doses of folinic acid may potentiate the angiogenesis activity of bevacizumab^13^. In the UK, however, bevacizumab is not currently routinely funded for treatment of colorectal cancer and so this is not of clinical relevance and there is a complete lack of randomised trial data to confirm the value of adding folinic acid in this setting.

Furthermore, the panel recommended that BOPA should encourage further research into the optimal schedule for dosing folinic acid in combination with fluorouracil. Adopting this statement (Median score: 1, IQR: 0):

> ***Recommendation 11:***
>
> *The British Oncology Pharmacy Association should support the recommendation of Amorim et al^1^ by stating that: international efforts should be made to implement randomised trials investigating the dosing, safety and efficacy of fluorouracil bolus and folinic acid.*

### Alternative treatments

The panel considered if they could give advice on the use of alternatives to fluorouracil to entirely avoid the risk of incompatibility. The panel considered several statements and only reached consensus on the following statement (Median score: 2, IQR: 0.75):

> ***Recommendation 12:***
>
> *In the absence of robust compatibility data, and where peer reviewed evidence of clinical efficacy is available, treatment with alternative drugs such as raltitrexed or capecitabine could be considered instead of fluorouracil, as they do not need to be administered with folinic acid.*

### Updating these guidelines

BOPA will review this practice guideline in the event that new information is brought to its attention. This may be in the form of published evidence, or further detailed case reports of incompatibility that suggest there is a pharmaceutically and clinically significant problem with co-infusion of calcium folinate and either oxaliplatin or irinotecan.

### Implementation of these guidelines

We believe these guidelines are simple, clear and pragmatic. We believe they could be quickly adopted in the UK, with revision of electronic chemotherapy ordering templates (Recommendations 1, 3, 4, 6 & 10), and internal aseptic preparation documentation (Recommendations 1 & 7). Those organisations which outsource supply of (unlicensed) folinic acid products for infusion should urgently review their unlicensed medicines policies and procedures to ensure that they meet regulatory requirements^14^ by accurately specifying full details of how all products are to be made and labelled. Organisations should also consider developing a standard operating procedure for investigation of incidents or review any existing procedures to align their clinical practice to this guideline (Recommendation 8).

### Resource implications

The aim of our recommendations are to minimise the possibility of adverse impact on the capacity of the UK’s already stretched chemotherapy day units, the significant overall adverse impact on patients’ valuable time and to mitigate risks to continuity of supply of calcium folinate products. Except for recommendation 5, we do not believe there should be significant financial impacts from implementation of this guidance. If they help to optimise dosing of folinic acid and hence eliminate reliance on off-contract procurement of licesnd products, they may even achieve signeifact cost benefits for some organisations.

### Monitoring and audit criteria

Recommendation 8 sets out arrangements for monitoring and investigation of any apparent incompatibility incidents. This should form the basis of monitoring of this guidance. Any future incidents which could be potentially related to this guidance should be reported to the relevant pharmacovigilance authority and to BOPA .

## Supporting information

Supplemental - Round 2 Voting Results

Supplemental - Round 3 Voting Results

Supplemental - Round 4 Voting Results

## Data Availability

All data produced in the present work are contained in the manuscript

## Acknowledgements

Nathan Ng (Sunnybrook), Anne Black (NHS Specialist Pharmacy Service), Lola Sibley (The Newcastle-upon-Tyne Hospitals NHS Foundation Trust), John Minshull (NHS Specialist Pharmacy Service).

## References

1. Amorim LC, Peixoto RD. Should we still be using bolus 5-FU prior to infusional regimens in gastrointestinal cancers? A practical review. Int Cancer Conf J. 2022;11(1):2–5. doi:10.1007/s13691-021-00526-7

2. Cheeseman SL, Joel SP, Chester JD, et al. A “modified de Gramont” regimen of fluorouracil, alone and with oxaliplatin, for advanced colorectal cancer. Br J Cancer. 2002;87(4):393–399. doi:10.1038/sj.bjc.6600467

3. Van Cutsem, E, Köhne, Ch, Hitre, E, et al. Cetuximab and chemotherapy as initial treatment for metastatic colorectal cancer. N Engl J Med. 2009;360(14). doi:10.1056/NEJMoa0805019

4. Etienne MC, Guillot T, Milano G. Critical factors for optimizing the 5-fluorouracil-folinic acid association in cancer chemotherapy. Ann Oncol. 1996;7(3):283–289. doi:10.1093/oxfordjournals.annonc.a010573

5. medac GmbH. Oxaliplatin medac 5 mg/ml concentrate for solution for infusion - Summary of Product Characteristics (SmPC) - (emc). Published April 3, 2019. Accessed September 12, 2019. https://www.medicines.org.uk/emc/product/7358/smpc

6. Summary of Product Characteristics: Calcium Folinate 10 mg/ml Solution for injection or infusion. Published online December 4, 2017. https://www.consilienthealth.co.uk//wp-content/uploads/2021/06/Calcium-Folinate-10-mgml-Solution-for-injection-or-infusion-SPC.pdf

7. Hayes MS, Ward MA, Slabaugh SL, Xu Y. Lessons from the leucovorin shortages between 2009 and 2012 in a medicare advantage population: where do we go from here? Am Health Drug Benefits. 2014;7(5):264–270.

8. Nasa P, Jain R, Juneja D. Delphi methodology in healthcare research: How to decide its appropriateness. World J Methodol. 2021;11(4):116–129. doi:10.5662/wjm.v11.i4.116

9. L Flay Charbonneau, Ivan Tyono, Jonathan Shloush, Nathan Ma. Stability and Compatibility of Oxaliplatin and Generic Medical Partners Inc. Leucovorin Formulation. J Oncol Pharm Pract. 2023;29(3S):14.

10. Polwart C. Calcium Folinate - BOPA Consensus Meeting. Published online 2023:671484 Bytes. doi:10.6084/M9.FIGSHARE.24146460.V1

11. Burki T. Systemic anti-cancer therapy treatment centres in Scotland at crisis point. Lancet Oncol. 2022;23(8):982–983. doi:10.1016/S1470-2045(22)00374-6

12. Walker SE, Law S, Puodziunas A. Simulation of Y-Site Compatibility of Irinotecan and Leucovorin at Room Temperature in 5% Dextrose in Water in 3 Different Containers. 2005;58(4).

13. Budai B, Nagy T, Láng I, Hitre E. The use of high dose d,l-leucovorin in first-line bevacizumab+mFOLFIRI treatment of patients with metastatic colorectal cancer may enhance the antiangiogenic effect of bevacizumab. Angiogenesis. 2013;16(1):113–121. doi:10.1007/s10456-012-9303-z

14. Williamson S, Beaney A, Polwart C. Guidance on managing the sourcing and supply of ready to administer chemotherapy doses for the NHS. Published online December 5, 2016. Accessed January 30, 2023. https://www.sps.nhs.uk/articles/guidance-on-managing-the-sourcing-and-supply-of-ready-to-administer-chemotheraphy-doses-for-the-nhs-yellow-cover/

