## Supplemental Information for "British Oncology Pharmacy Association Delphi Consensus Guidelines: co-infusion of trometamol-containing calcium folinate (Leucovorin) with Systemic Anti-Cancer Treatments"

Materials

### Membership of the original round table discussion

- Tim Root FRPharmS, Assistant Head of NHS Specialist Pharmacy Service. Former chair of British Oncology Pharmacy Association. (Discussion Chair)
- Ian Purcell, Advanced Pharmacy Practitioner/Team Leader – Oncology, Nottingham University Hospitals
- Ibrahim Al-Modaris, Principal Pharmacist, Haematology & Oncology, Chesterfield Royal Hospitals
- Mohammed Patel, Lead Oncology Pharmacist, Bradford Teaching Hospitals
- Calum Polwart MRPharmS (Consultant), Consultant Pharmacist, South Tees Hospitals
- Bruce Burnett, Research Officer & Data Scientist, Swansea University (at time of discussion),
- Rhiannon Walters-Davies, Chair of the British Oncology Pharmacy Association Guidelines and Publications Sub-Committee and Principal Pharmacist at Velindre Cancer Centre, Wales (at time of discussion)..
- John Minshull, Professional Lead for Medicines Advice, NHS Specialist Pharmacy Service

### Membership of the voting panel (Round 2 onwards)

| **Name** | **Organisation** |
| --- | --- |
| Ana Coelho^1^ | Nuffield Health^a,b^ |
| Andrew Craig^1^ | Stockport NHS Foundation Trust^a,b^ |
| David Barber^1^ | Lancashire Teaching Hospitals NHS Trust^a,b^ |
| Bill Wetherill^1^ | South Tees Hospitals NHS Foundation Trust^d^ |
| Calum Polwart^1^ | South Tees Hospitals NHS Foundation Trust^a,b,d^ |
| Caroline Clapham^2^ | Bedford Hospitals NHS Trust^a,b^ |
| Chauhan Rena^1^ | Guys’ & St Thomas’ NHS Foundation Trust^b,c^ |
| Chloë Waterson^1^ | York and Scarborough Teaching Hospital NHS Foundation Trust^a,b,d^ |
| Gareth Tyrrell^1^ | NHS Wales Shared Services Partnership^a,b,d,e^ |
| Ibrahim Al-Modaris^1^ | Sheffield Teaching Hospitals NHS Foundation Trust^a,b^ |
| Janitha Vasavan^1^ | Chelsea & Westminster Hospital NHS Foundation Trust^b,d^ |
| Kate Reeves^1^ | Royal Berkshire NHS Foundation Trust^h^ |
| Nathan Ma^1^ | Sunnybrook Hospital, Canada^g^ |
| Ruth Orchard^1^ | Leeds Teaching Hospitals NHS Trust^a,b^ |
| Shah Aneri^1^ | Guys’ & St Thomas’ NHS Foundation Trust^a,b^ |
| Simon Jenkinson^1^ | Royal Free London NHS Foundation Trust^b^ |
| Songul Tezcan^1^ | Marmara University, Turkey^c,d,i^ |
| Stephen Wardell^1^ | The Christie NHS Foundation Trust^a^ |
| Tim Root^1^ | NHS Specialised Pharmacy Services^c,d,f^ |

Professional Group: ^1^Pharmacist, ^2^Registered Nurse

Areas of Expertise: ^a^Clinical Pharmacy – Upper GI [11], ^b^Clinical Pharmacy – Lower GI [12], ^c^Oncology Pharmacy [3], ^d^Aseptic Pharmacy [7], ^e^Procurement[1], ^f^Specialised Support Services[1], ^g^Quality Assurance[1], ^h^Chemotherapy Nurse[1], ^i^Academia[1]

### Number of votes for each round

| **Delphi Round** | **Voters** |
| --- | --- |
| Round 2 | 18 |
| Round 3 | 13 |
| Round 4 | 14 |

### Questions posed

|  | **Question** | **Delphi Round** | | | **Recommendation** |
| --- | --- | --- | --- | --- | --- |
|  |  | 2 | 3 | 4 |  |
| 1A | Oxaliplatin appears to be safe when co-administered through the same intravenous line as calcium folinate which contains trometamol when given in concentrations between 0.2mg/mL and 0.7mg/mL of oxaliplatin and 0.35mg/mL and 1.54mg/mL of calcium folinate, based on the evidence from Flay et al. | Y | Y | Y |  |
| 1B | Oxaliplatin appears to be safe when co-administered through the same intravenous line as calcium folinate which contains trometamol when given in concentrations between 0.2mg/mL and 0.7mg/mL of oxaliplatin and 0.35mg/mL and 1.54mg/mL of calcium folinate, based on the evidence from Flay et al. Only after peer review of the Flay et al study by a independent Quality Assurance expert (for example from an NHS Quality Assurance Service). | N | Y | N |  |
| 1C | The anti-cancer activity of oxaliplatin does not appear to be reduced and it can be safely co-administered through the same intravenous line as calcium folinate which contains trometamol when given in concentrations between 0.2mg/mL and 0.7mg/mL of oxaliplatin and 0.35mg/mL and 1.54mg/mL of calcium folinate, based on the evidence from Flay et al. | N | N | Y |  |
| 1D | The stability of oxaliplatin does not appear to be changed and it can be safely co-administered through the same intravenous line as calcium folinate which contains trometamol when given in concentrations between 0.2mg/mL and 0.7mg/mL of oxaliplatin and 0.35mg/mL and 1.54mg/mL of calcium folinate, based on the evidence from Flay et al. | N | N | Y | 1 |
| 2A | There is no data to confirm the safety of co-administration of irinotecan with trometamol containing calcium folinate, and therefore co-administration through a y-site is not recommended. | Y | Y | N | 4 |
| 2B | There is no data to confirm the safety of co-administration of irinotecan with trometamol containing calcium folinate, and therefore co-administration through a y-site could be recommended. | N | Y | N |  |
| 2C | In the absence of any data or reports of adverse reactions which suggest reduced clinical activity after co-administration of irinotecan with trometamol containing calcium folinate through a y-site suggest that it is safe | N | Y | N |  |
| 3A | For administration with irinotecan containing regimens, such as FOLFIRI or FOLFIRINOX, irinotecan could be given over 30 – 90 minutes followed by a 30 – 90 minute infusion of calcium folinate allowing completion of administration within 2 hours. | Y | Y | N | 6 |
| 3B | For administration with irinotecan containing regimens, such as FOLFIRI or FOLFIRINOX, irinotecan could be given over 30 – 90 minutes followed by a 180 minute infusion of calcium folinate allowing completion of administration within 3.5 hours. | N | Y | N |  |
| 3C | For administration with irinotecan containing regimens, such as FOLFIRI or FOLFIRINOX, irinotecan could be given over 30 – 60 minutes followed by a 30 – 60 minute infusion of calcium folinate allowing completion of administration within 2 hours. | N | Y | N |  |
| 3D | 3D - For administration with irinotecan containing regimens, such as FOLFIRI or FOLFIRINOX, irinotecan could be given over up to 90 minutes followed by a 30 minute infusion of calcium folinate allowing completion of administration within 2 hours. | N | Y | N |  |
| 4A | There is a lack of clear data supporting different doses of calcium folinate in de Gramont derived protocols. Calcium folinate frequently experiences stock shortages, and therefore use of higher doses (e.g. 400mg/m2) without clear evidence of benefit may cause harm to other patients in the future. | Y | Y | N |  |
| 4B | There is a lack of clear data supporting different doses of calcium folinate in de Gramont derived protocols. Calcium folinate frequently experiences stock shortages, and therefore use of higher doses (e.g. 400mg/m2) of folinic acid without clear evidence of benefit may indirectly compromise the treatment of other patients if folinic acid isn't immediately available to continue their treatment. | N | Y | N | 9 |
| 5A | Use of a consistent fixed dose of calcium folinate (such as 350mg) may enable manufacturers to optimise the supply chain to the correct vial size and is encouraged. | Y | Y | N | 10 |
| 5B | Use of a consistent fixed dose of calcium folinate (such as 300mg) may enable manufacturers to optimise the supply chain to the correct vial size and is encouraged. | N | Y | N |  |
| 5C | Use of a consistent fixed dose of calcium folinate (such as 350mg) may enable manufacturers to optimise the supply chain to the correct vial size and is encouraged. In the event of a supply shortage, 300mg is a suitable fixed dose. | N | Y | N |  |
| 6A | The British Oncology Pharmacy Association should support the recommendation of Amorim et al by stating that: international efforts should be made to implement randomised trials investigating the safety and efficacy of fluorouracil bolus and folinic acid. | Y | Y | N | 11 |
| 7A | Any calcium folinate product being supplied either from the hospital pharmacy as a pre-prepared product, or sourced from another supplier as a pre-prepared product should clearly state if it contains trometamol as an excipient. For third party supply arrangements, the purchasing pharmacy must ensure their product specifications are adequate to require this. | Y | Y | N |  |
| 7B | Any calcium folinate product being supplied either from the hospital pharmacy as a pre-prepared product, or sourced from another supplier as a pre-prepared product should clearly state if it contains trometamol as an excipient, giving an indication of concentration. For third party supply arrangements, the purchasing pharmacy must ensure their product specifications are adequate to require this. | N | Y | N | 7 |
| 8A | Trusts should ensure there are adequate policies and systems in-place to ensure glucose 5% is used as a flush with oxaliplatin, and sodium chloride 0.9% is not used as a diluent with calcium folinate when given with oxaliplatin. | Y | N | N | 3 |
| 9A | Any incidents of apparent incompatibility must be thoroughly documented, investigated and reported. This must include - batch numbers of raw ingredients, final concentrations, diluents and flushes. | Y | Y | N |  |
| 9B | Any incidents of apparent incompatibility must be thoroughly documented, investigated and reported. This must include - manufacturer details, batch numbers and expiries of raw ingredients, final concentrations, diluents and flushes, as well as storage and expiry date of infusion bag. | N | Y | N | 8 |
| 10A | In the absence of robust compatibility data, raltitrexed or capecitabine should be considered instead of fluorouracil as they do not need to be administered with folinic acid. | N | Y | Y |  |
| 10B | In the absence of robust compatibility data, and where peer reviewed evidence of clinical efficacy is available, treatment with alternative drugs such as raltitrexed or capecitabine should be considered instead of fluorouracil, as they do not need to be administered with folinic acid. | N | N | Y |  |
| 10C | In the absence of robust compatibility data, and where peer reviewed evidence of clinical efficacy is available, treatment with alternative drugs such as raltitrexed or capecitabine could be considered instead of fluorouracil, as they do not need to be administered with folinic acid. | N | N | Y | 12 |
| 10D | In the absence of robust compatibility data, treatment with raltitrexed or capecitabine could be considered as an alternative to fluorouracil as they do not need to be administered with folinic acid. | N | N | Y |  |
| 11A | A compatibility study of irinotecan and trometamol containing calcium folinate should be commissioned. | N | Y | N | 5 |
| 12A | The Flay et al study of compatibility only demonstrated compatibility when oxaliplatin and calcium folinate were mixed for a period of 3 hours. When oxaliplatin and calcium folinate are infused through a y-site the contact time in a single lumen line when administered through a y-site is short, even if the infusion duration is prolonged. Therefore no time limit, for reasons of compatibility, is required when co-infusing oxaliplatin and calcium folinate. | N | N | Y | 2 |
| 12B | The Flay et al study of compatibility only demonstrated compatibility when oxaliplatin and calcium folinate were mixed for a period of 3 hours. When oxaliplatin and calcium folinate are infused through a single lumen line using a y-site, the calcium folinate should normally be infused over the final 2 hours of the planned oxaliplatin infusion. | N | N | Y |  |
| 13 | Do you agree with the grammatical amendments described below  Previously agreed statements should have minor grammatical improvements such as:   - "...trometamol containing..." being amended to "...trometamol-containing..." (Alters statements 2A, and 11) - "...irinotecan containing..." being amended to "...irinotecan-containing..." (Alters statements 3A) - If we accept compatibility with oxaliplatin, compatibility with irinotecan is only relevant when not being administered in a regime also containing oxaliplatin. Therefore FOLFIRINOX can be removed from statement 3. - statements regarding y-sites should specifically state "through a single lumen line" (Alters statements 2A) - statement 7 regarding products specification should state "ensure their product specifications are adequate to require this information to be provided by the supplier" - "Trusts" should be amended to "Hospitals" to be inclusive of all providers in the UK both in the NHS and private sectors (Alters statement 8) - "Raw materials" should be amended to "licensed starting materials", and "storage" should be amended to "storage conditions". (Alters 9B) - The second sentence of statement 9B should begin "The report" rather than "This". (Alters 9B) - "Flushes" should state flushes used (Alters statement 9B) | N | N | Y |  |
