## Supplemental - Round 2 Voting Results for "British Oncology Pharmacy Association Delphi Consensus Guidelines: co-infusion of trometamol-containing calcium folinate (Leucovorin) with Systemic Anti-Cancer Treatments"

| Statement | Median | IQR | Consensus |
|---|---|---|---|
| Oxaliplatin appears to be safe when co-administered through the same intravenous line as calcium folinate which contains trometamol when given in concentrations between 0.2mg/mL and 0.7mg/mL of ox... | 1 | 1 | Yes - Agree |
| There is no data to confirm the safety of co-administration of irinotecan with trometamol containing calcium folinate, and therefore co-administration through a y-site is not recommended. | 2 | 1 | Yes - Agree |
| For administration with irinotecan containing regimens, such as FOLFIRI or FOLFIRINOX, irinotecan could be given over 30 – 90 minutes followed by a 30 – 90 minute infusion of calcium folinate allo... | 2 | 1 | Yes - Agree |
| There is a lack of clear data supporting different doses of calcium folinate in de Gramont derived protocols. Calcium folinate frequently experiences stock shortages, and therefore use of higher d... | 2 | 0.75 | Yes - Agree |
| Use of a consistent fixed dose of calcium folinate (such as 350mg) may enable manufacturers to optimise the supply chain to the correct vial size and is encouraged. | 1 | 1 | Yes - Agree |
| The British Oncology Pharmacy Association should support the recommendation of Amorim et al by stating that: international efforts should be made to implement randomised trials investigating the ... | 1.5 | 1 | Yes - Agree |
| Any calcium folinate product being supplied either from the hospital pharmacy as a pre-prepared product, or sourced from another supplier as a pre-prepared product should clearly state if it conta... | 1 | 0 | Yes - Agree |
| Trusts should ensure there are adequate policies and systems in-place to ensure glucose 5% is used as a flush with oxaliplatin, and sodium chloride 0.9% is not used as a diluent with calcium folin... | 1 | 0 | Yes - Agree |
| Any incidents of apparent incompatibility must be thoroughly documented, investigated and reported. This must include - batch numbers of raw ingredients, final concentrations, diluents and flushes. | 1 | 0 | Yes - Agree |
