## Supplemental - Round 3 Voting Results for "British Oncology Pharmacy Association Delphi Consensus Guidelines: co-infusion of trometamol-containing calcium folinate (Leucovorin) with Systemic Anti-Cancer Treatments"

| Statement | Median | IQR | Consensus |
|---|---|---|---|
| Having reached consensus on all the statements, do you feel we should still try and improve the statements? | NA | NA | No |
| 1A Oxaliplatin appears to be safe when co-administered through the same intravenous line as calcium folinate which contains trometamol when given in concentrations between 0.2mg/mL and 0.7mg/mL of... | 2 | 0 | Yes - Agree |
| 1B - Oxaliplatin appears to be safe when co-administered through the same intravenous line as calcium folinate which contains trometamol when given in concentrations between 0.2mg/mL and 0.7mg/mL ... | 1 | 0 | Yes - Agree |
| 2A - There is no data to confirm the safety of co-administration of irinotecan with trometamol containing calcium folinate, and therefore co-administration through a y-site is not recommended. | 2 | 1.5 | Yes - Agree |
| 2B - There is no data to confirm the safety of co-administration of irinotecan with trometamol containing calcium folinate, and therefore co-administration through a y-site could be recommended. | 4 | 0.75 | Yes - Disagree |
| 2C - In the absence of any data or reports of adverse reactions which suggest reduced clinical activity after co-administration of irinotecan with trometamol containing calcium folinate through a ... | 4 | 2 | No |
| 3A - For administration with irinotecan containing regimens, such as FOLFIRI or FOLFIRINOX, irinotecan could be given over 30 – 90 minutes followed by a 30 – 90 minute infusion of calcium folinate... | 2 | 0.5 | Yes - Agree |
| 3B - For administration with irinotecan containing regimens, such as FOLFIRI or FOLFIRINOX, irinotecan could be given over 30 – 90 minutes followed by a 180 minute infusion of calcium folinate al... | 4 | 0.75 | Yes - Disagree |
| 3C - For administration with irinotecan containing regimens, such as FOLFIRI or FOLFIRINOX, irinotecan could be given over 30 – 60 minutes followed by a 30 – 60 minute infusion of calcium folinate... | 2 | 0 | Yes - Agree |
| 3D - For administration with irinotecan containing regimens, such as FOLFIRI or FOLFIRINOX, irinotecan could be given over up to 90 minutes followed by a 30 minute infusion of calcium folinate al... | 2 | 2.75 | No |
| 4A - There is a lack of clear data supporting different doses of calcium folinate in de Gramont derived protocols. Calcium folinate frequently experiences stock shortages, and therefore use of hig... | 2 | 0 | Yes - Agree |
| 4B - There is a lack of clear data supporting different doses of calcium folinate in de Gramont derived protocols. Calcium folinate | 1 | 0 | Yes - Agree |

| Statement | Median | IQR | Consensus |
|---|---|---|---|
| frequently experiences stock shortages, and therefore use of ... | | | |
| 5A - Use of a consistent fixed dose of calcium folinate (such as 350mg) may enable manufacturers to optimise the supply chain to the correct vial size and is encouraged. | 1 | 1 | Yes - Agree |
| 5B - Use of a consistent fixed dose of calcium folinate (such as 300mg) may enable manufacturers to optimise the supply chain to the correct vial size and is encouraged. | 2 | 1.75 | No |
| 5C - Use of a consistent fixed dose of calcium folinate (such as 350mg) may enable manufacturers to optimise the supply chain to the correct vial size and is encouraged. In the event of a supply ... | 2 | 1 | Yes - Agree |
| 6A - The British Oncology Pharmacy Association should support the recommendation of Amorim et al by stating that: international efforts should be made to implement randomised trials investigating... | 2 | 0 | Yes - Agree |
| 6B - The British Oncology Pharmacy Association should support the recommendation of Amorim et al by stating that: international efforts should be made to implement randomised trials investigating ... | 1 | 0 | Yes - Agree |
| 7A - Any calcium folinate product being supplied either from the hospital pharmacy as a pre-prepared product, or sourced from another supplier as a pre-prepared product should clearly state if it ... | 2 | 0 | Yes - Agree |
| 7B - Any calcium folinate product being supplied either from the hospital pharmacy as a pre-prepared product, or sourced from another supplier as a pre-prepared product should clearly state if it ... | 1 | 0 | Yes - Agree |
| 9A - Any incidents of apparent incompatibility must be thoroughly documented, investigated and reported. This must include - batch numbers of raw ingredients, final concentrations, diluents and f... | 2 | 0 | Yes - Agree |
| 9B - Any incidents of apparent incompatibility must be thoroughly documented, investigated and reported. This must include - manufacturer details, batch numbers and expiries of raw ingredients, f... | 1 | 0 | Yes - Agree |
| NEW: In the absence of robust compatibility data, raltitrexed or capecitabine should be considered instead fluorouracil as they do not need to be administered with folinic acid. | 3 | 2 | No |
| NEW: A compatibility study of irinotecan and trometamol containing calcium folinate should be commissioned. | 1 | 1 | Yes - Agree |

| Statement | Median | IQR | Consensus |
| --- | --- | --- | --- |
| Do you have any conflicts of interest that need to be declared: | NA | NA | No |
