## Supplemental - Round 4 Voting Results for "British Oncology Pharmacy Association Delphi Consensus Guidelines: co-infusion of trometamol-containing calcium folinate (Leucovorin) with Systemic Anti-Cancer Treatments"

| Statement | Median | IQR | Consensus |
|---|---|---|---|
| Do you agree with the gramatical ammendments described below Previously agreed statements should have minor grammatical improvements such as: "...trometamol containing..." being amended to "...... | 1 | 1 | Yes - Agree |
| 1A Oxaliplatin appears to be safe when co-administered through the same intravenous line as calcium folinate which contains trometamol when given in concentrations between 0.2mg/mL and 0.7mg/mL of... | 2 | 0.75 | Yes - Agree |
| 1C The anti-cancer activity of oxaliplatin does not appear to be reduced and it can be safely co-administered through the same intravenous line as calcium folinate which contains trometamol when g... | 3 | 2 | No |
| 1D The stability of oxaliplatin does not appear to be changed and it can be safely co-administered through the same intravenous line as calcium folinate which contains trometamol when given in con... | 1 | 1 | Yes - Agree |
| 12A The Flay et al study of compatability only demonstrated compatability when oxaliplatin and calcium folinate were mixed for a period of 3 hours. When oxaliplatin and calcium folinate are infu... | 2 | 0.75 | Yes - Agree |
| 12B The Flay et al study of compatability only demonstrated compatability when oxaliplatin and calcium folinate were mixed for a period of 3 hours. When oxaliplatin and calcium folinate are infus... | 2 | 1.75 | No |
| 10A In the absence of robust compatibility data, raltitrexed or capecitabine should be considered instead fluorouracil as they do not need to be administered with folinic acid. | 3 | 1 | No |
| 10B In the absence of robust compatibility data, and where peer reviewed evidence of clinical efficacy is available, treatment with alternative drugs such as raltitrexed or capecitabine should be ... | 3 | 1.75 | No |
| 10C In the absence of robust compatibility data, and where peer reviewed evidence of clinical efficacy is available, treatment with alternative drugs such as raltitrexed or capecitabine could be c... | 2 | 0.75 | Yes - Agree |
| 10D In the absence of robust compatibility data, treatment with raltitrexed or capecitabine could be considered as an alternative to fluorouracil as they do not need to be administered with folini... | 3 | 1.75 | No |
| Do you have any conflicts of interest that need to be declared: | NA | NA | No |
